# Medical Students Perception of Anatomage: A 3D Interactive (Virtual) Anatomy Dissection Table

**DOI:** 10.1101/2022.04.25.22274178

**Authors:** A. Elizabeth Memudu, Idaguko C. Anna, M. Oluwatosin Gabriel, Augustine Oviosun, W. Barinem Vidona, A. Amoo Odetola, S. Ehizokhale Ehehba, O. Abimbola Ebeye, A. Obioma Nwaopara, N. Willi Dare, Akinyinka O. Olusegun

## Abstract

**Introduction:** The rising number of Medical Schools and the increasing demand for cadavers, amid its scarcity, has prompted the search for alternatives in Anatomy Education. This study assessed students’ thought of the use of Anatomage as an Anatomy teaching and learning tool in medical school.

**Methods:** A five-point scale questionnaire with a free hand comment section was completed by 50 medical students exposed to the use of Anatomage alongside the traditional cadaveric dissection for 2 academic sessions.

**Results:** Our results findings showed that there were preference pattern variations in the use of the Anatomage for various fields of anatomy such as Gross Anatomy (48%), Histology (46%), and Neuroanatomy (2%) respectively. Furthermore, 66% opined that Anatomage and Cadaveric dissection should be complementary in teaching and learning anatomy. However, been satisfied with Anatomage was 76% (52% completely and 24% generally agreed respectively), while Anatomage increasing their interest in Anatomy was 66% (40% completely agreed and 26% generally agreed) and 74% (40% completely agreed and 34% generally agreed) of learning outcomes been achieved using Anatomage. Also, 68% stated that the micrographs were well displayed for histology teaching. Overall, 60% of the students agreed that Anatomage should be encouraged in teaching and learning Anatomy, along with other teaching aids.

**Discussion:** Anatomage increased students’ interest in Anatomy as its 3D-image display enabled better visualization of relevant anatomical structures. Anatomage has the potential to be a beneficial supplement to standard learning methods in the acquisition of 3D anatomy information.

## Introduction

Anatomy is the fundamental subject in basic Medical Science Education [1, 2]. The significance of the methods of teaching anatomy and its competence in high-quality health care and medical training curriculum has been debated over the years [3, 4, 5]. Several studies on teaching Human Anatomy have already compared new teaching methods to traditional ones [6, 7]. Traditionally, teaching and studying of Anatomy are achieved through Cadaveric dissection [8]. This method is mostly adopted in most Medical Schools’ Curricula [9]. The act of dissection involves the exposure and description of internal body organs and structures using deceased animal or human bodies [10, 11, 12]. Over the years, however, new and more interactive methods to teaching anatomy have emerged and are used by students and anatomy educators [13]. The developmental intensity in technologies over the years has profoundly affected the teaching of Anatomy in the last decade and there is much debate about the suitable methods of delivering anatomical knowledge to students; hence the resolve by many medical tutors to use the three-dimensional display equipment [14]. The explosion of technologies during the last few decades has brought anatomical education into a new world [15, 4]. Some of these emerging technologies allow virtual cadaver dissections by applying the principle of simulator soft-wares, these include the Zygote, Anatomage, and Sectra tables [16, 17, 18].

The Anatomage Table was developed by a 3-D Medical Technology Company located in San José (California) in conjunction with Stanford University’s Clinical Anatomy Division. Historically, it was conceptualized due to the ethical and health issues around cadaveric dissection and the need to key into the technological ‘wind of change’ triggering the emergence of several innovative tools, which enables virtual anatomy education.

The design and development began in 2004 when the first volume construction software for the dental market was developed; with its flagship product being the In vivo-Dental [19, 20]. Then in 2011, the Anatomage Table was released as a platform to present Anatomy on a life-size scale, where the same technology of In vivo-Dental Software was adopted and the use of cone-beam of Computerized Tomography (CT) and Magnetic Resonance Imaging (MRI) was applied on the scan files of human images. These were rendered into 3D to create the Anatomage Table [21, 22].

In terms of functionality, the Anatomage is a cutting-edge anatomy visualization system for anatomy instruction and an interface that allows students to explore life-size anatomy on an interactive 3D table [23]. The technology allows virtual dissection and reconstruction of the images. Presently, many of the world’s premier medical schools and institutions have now adopted the Anatomage [18]. However, discussions about the qualities and limitations of alternative teaching resources like the Anatomage are ongoing [24], particularly in this era of continuous developments in computer/mobile applications and evolution in the use of interactive technology in medical sciences [25, 26].

As a result, the purpose of this study was to find out what third-year medical students at Edo State University in Uzairue, Edo State, Nigeria, thought about the use of anatomage in teaching and studying anatomy.

## Methods

### Ethical Consideration

Approval for the study was obtained from the Basic Medical Ethics Committee, Edo State University Uzairue. All the subjects were informed of the purpose of the research and their consent sort before the commencement of the study. They consented to participate in the study and signed informed consent forms.

### Study design

A questionnaire and focus discussion-based exploratory study was designed to assess the comparative impact of Anatomage Table (version 7.00) usage in the teaching of anatomy to 300L (third year) Medical Students at Edo State University, Uzairue, Edo State, Nigeria.

### Area of study

The study took place at Edo State University Uzairue (EDSU), within Iyamho (7.2193° N, 6.3344° E), in Etsako West Local Government Area of Edo State, Nigeria. The University was approved to commence academic activities in 2016, by the National University Commission (NUC), Nigeria, as the 41st State-owned University in Nigeria.

### Study population

A total of 50 students, of which 20 were males (40%) and 30 were females (60%) were enrolled in the study. They were third-year medical students of the College of Medicine, Edo State University Uzairue, Edo State.

### Inclusion and Exclusion Criteria

The essential inclusion attributes for this study were that the students must be medical students of the Edo State University Uzairue and already exposed to the use of Anatomage in the course of teaching and learning.

### Activities with the Anatomage

Before data collection, the Anatomage table was used in 2 academic sessions and the courses taught were Gross Anatomy (upper and lower limbs, thorax and abdomen, pelvis and perineum, head and neck), Histology (basic and systemic), Neuroanatomy, Embryology (general and systemic). The teachers teaching these courses incorporated the Anatomage as an interactive educational tool into their classroom sessions.

The classroom activities included 1) viewing anatomy structures in 3D; 2) dissections; 3) viewing and presentation of pathological features, and 4) using the Anatomage table quiz/drill and practice functions for class activities.

### Preference patterns of respondents on the use of the Anatomage

Using a 5-point Likert scale questionnaire (The responses were categorized as follows: No experience of the topic = 1; I do not agree = 2; I slightly agree = 3; I generally agree = 4; I completely agree = 5.

The focus variables included the year of study, gender, perceptions about the Anatomage, and the associated reasons for liking or not liking anatomy, advantages of learning gross anatomy through the use of Anatomage; would they prefer other techniques of learning anatomy. Furthermore, in assessing the preference patterns of students on the use of Anatomage, the following sub-themes were investigated as part of the research questions in the questionnaire: the aspect of Anatomy best taught with the Anatomage (histology, gross anatomy, neuroanatomy, embryology, and no reply) and Anatomy teaching aid preference (only Anatomage, Anatomage with cadaver, cadaver only, models, animations and videos, no reply).

## Data Analysis

The data obtained from the questionnaires were analyzed using the Graphpad Prism 6 versions. Descriptive statistics in the form of percentages were worked out and recorded using Microsoft Excel Office Suite 2016.

## Results

The result shows that most of the respondents preferred the Anatomage for gross anatomy (48%), followed by histology (46%). However, none of the students considered it as a preference for teaching embryology as shown in figure 1.

**Figure 1:**
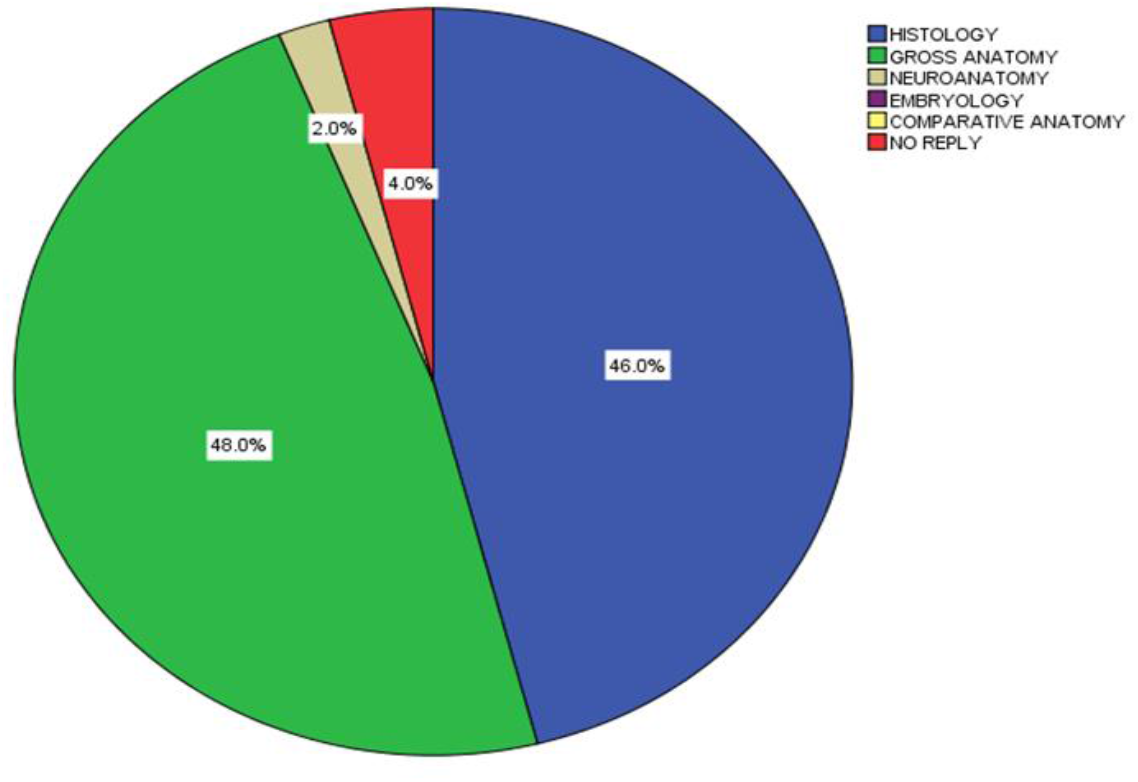
A pie chart showing the aspect of Anatomy best taught with the Anatomage table.

Students’ preference for teaching techniques in Anatomy showed that none of them preferred Anatomage alone and 2% preferred cadaver dissection alone. However, majority of respondents (66%) selected the combination of Anatomage and cadaver dissection as their preferred teaching technique. This result also provided evidence that the use of animations and videos remain relevant and effective in the teaching of Anatomy with relatively fewer respondents preferring it for the teaching of Anatomy (figure 2).

**Figure 2:**
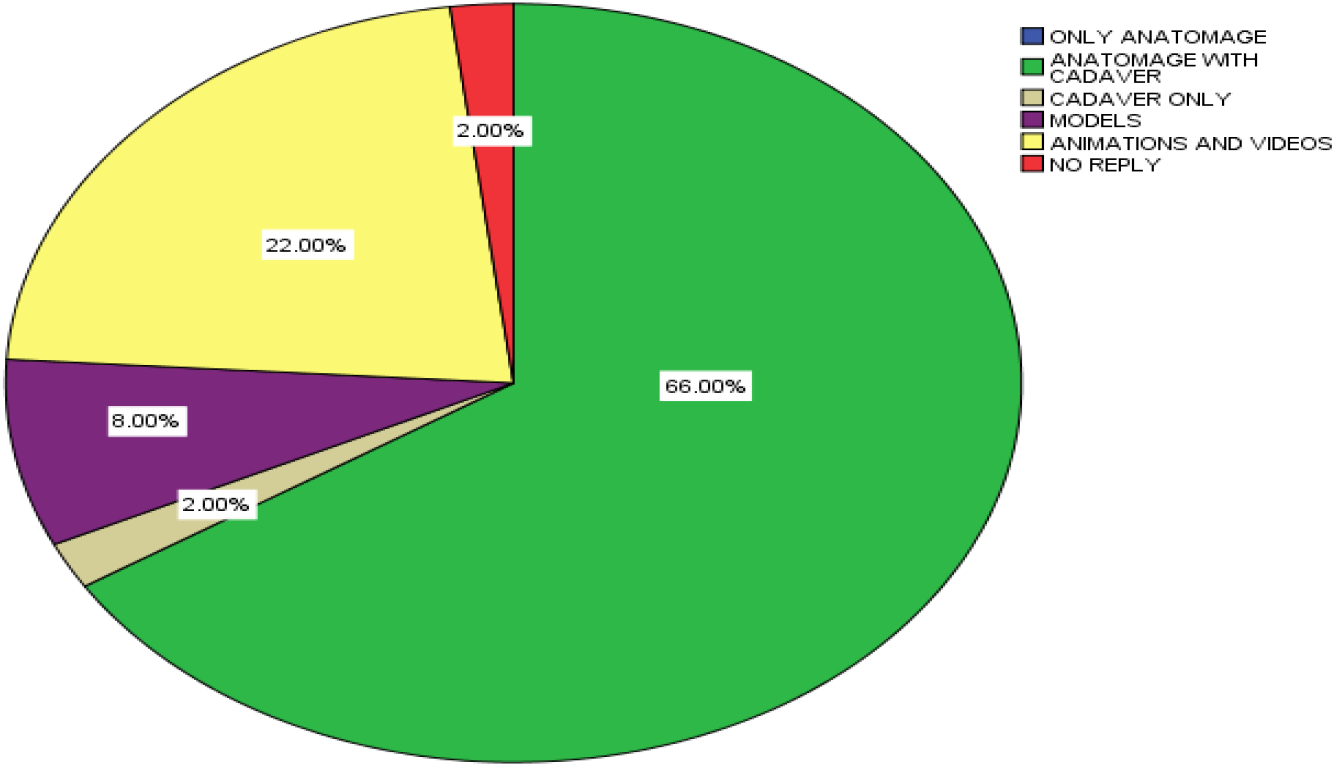
A pie-chart illustrating Comparative Anatomy teaching techniques surveyed.

Satisfied with Anatomage as a teaching tool; 52 % completely agreed and 24% generally agreed, while 40% completely agreed and 26% generally agreed that the use of Anatomage increased their interest in learning Anatomy. Also, 40 % completely agreed and 34% generally agreed that the learning outcome in using Anatomage was achieved as shown in figure 3.

**Figure 3:**
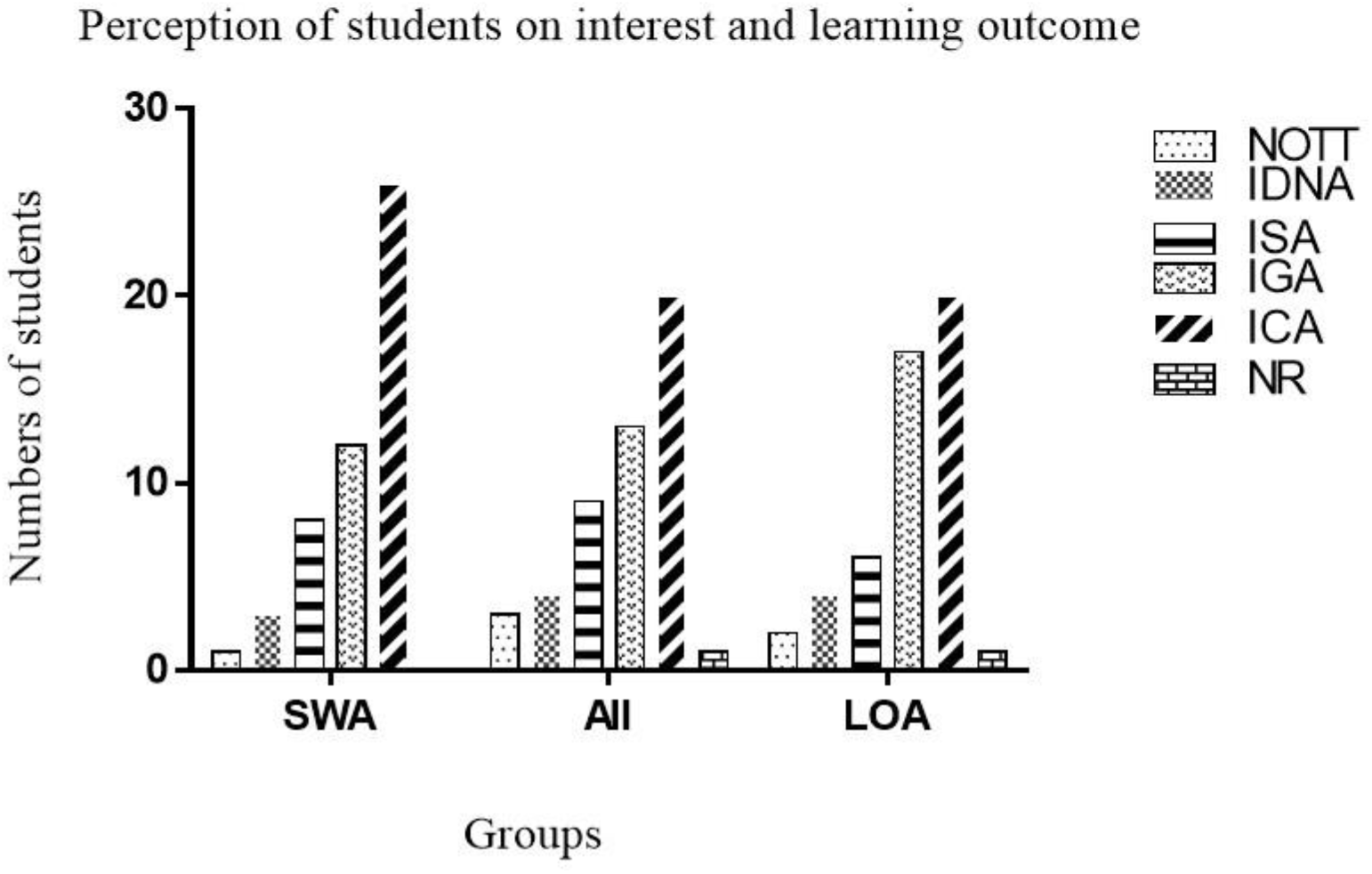
Perception of students on interest and learning outcome using the Anatomage. **Keys:** NOTT = No experience on the topic; IDNA = I do not agree; ISA = I slightly agree; IGA = I generally agree; ICA = I completely agree; NR = No reply. SWA: Satisfied With Anatomage, AII: Anatomage Increased Interest, LOA: Learning Outcome Achieved

Table i showed that 68% agreed that viewing and presenting histology slides on the *Anatomage* was better. While 60% was of the opinion that the use of *Anatomage* Table in the teaching of Anatomy should be encouraged.

**Table i:**
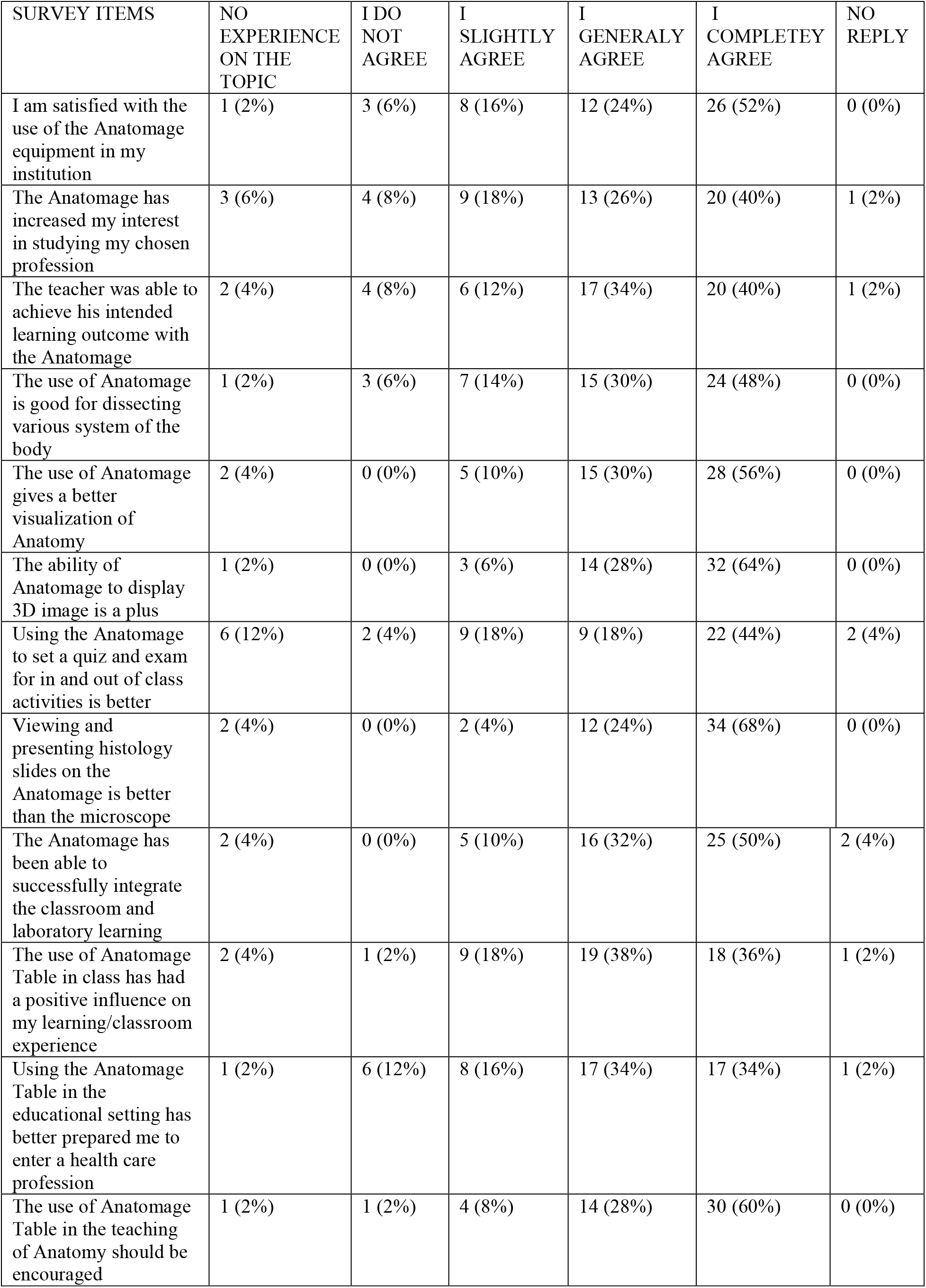
Responses on the use of the Anatomage in achieving learning outcomes and enhancing students’ interest in the study of Anatomy

## Discussion

Learning anatomy via 3D visual models is becoming increasingly prevalent with the use of virtual dissections [21], [27] reported that the students’ scores in Anatomy improved after they were taught using Anatomage as compared to their scores before exposure. Another report by [28] described the Anatomage table, as an effective Anatomy learning tool, giving that it increased students’ scores by 27.3%. [29] had earlier observed that students find the ability of Anatomage table to rotate structures very good and this has helped them while studying organ relationships. However, the tutors were not impressed with its technical issues; though they agree it is a good investment in anatomy education. But generally, there seems to be an increasing consensus that the Anatomage Table enhances learning by facilitating the hands-on and interactive approach to studying anatomy [27].

Our finding that most of our students were satisfied with the use of Anatomage in Anatomy education, is in line with the reports by [30] that students’ interest and satisfaction increases with the use of Anatomage. Furthermore, the significance of 3D in learning and teaching Anatomy has been highlighted by [31, 32, 33].

The technology is currently in use by most of the leading Medical Schools around the world [34] and its inclusion has helped increase overall students’ interest in Anatomy; as similarly observed in this study. On better visualization of Anatomic structures, it is noteworthy, to state that the reported better/ enhanced visualization of Anatomic structure is linked to the 3D imaging technology adopted in the production of the Anatomage. [33] had earlier described the significance of 3D/ spatial visualization of an anatomical structure as well as spatial relationship of anatomical structures to surrounding structures by students during teaching and learning of Anatomy.

This approach in learning Anatomy cannot be acquired using anatomical textbooks and atlas since they display the 2-D static anatomical illustration as compared to the 3D anatomical display [35]. Additionally, medical students need to have the ability to manipulate structures in 3D to appreciate their anatomy - visual-spatial learning [36, 37, 38, 31]. [39] have noted the use of digital 3D anatomic models such as anatomical visualization systems as an effective tool in enhancing teaching, learning and retention in medical schools. Available facts indicate also that medical students tend to prefer the use of 3D models to 2D images in Anatomy education [40].

Associated with the 3D model approach are the abilities to manipulate the model and create unlimited depictions of anatomical structures using technology, as well as enhanced spatial knowledge for learners, increased opportunity for collaborative and experiential learning, and increased motivation and commitment for learners [41].

More so, reports are acknowledging that 3D-Visualization improves learning outcomes and one’s effectiveness in appreciating anatomical structures’ spatial relationships [42, 43, 44]. However, it has been suggested that 3D methods of learning Anatomy can effectively be used to complement traditional teaching methods [45, 46]. In comparative terms, however, [47] have observed that students exposed to 3D/Virtual dissection (or Teaching method of Anatomy) performed better than those exposed to 2D (Anatomy Textbooks and Colored Atlas), though no significant difference was observed between those exposed to 3D and traditional dissection.

It is deducible from our results that the Anatomage successfully integrated classroom and laboratory learning as acknowledged by [48, 23, 49].

Above all, the students’ conviction that the use of Anatomage is used alongside other Anatomy teaching approach such as Cadaver dissection is in line with the findings by [23, 2], as sixty-six percent (66%) of the students wants it used along with the cadaveric dissection. This system will not only assist the students to learn anatomical details but also provide them the opportunity to appreciate 3D structures, which, according to Thompson [50], enhanced students’ capacity ability to learn complex anatomy structures better and faster than using only the traditional methods.

## Conclusion

Anatomage can play a significant role in the attaining 3D anatomy knowledge and promises to be a useful adjunct to traditional learning modalities. However, the complimentary use of the virtual Anatomage dissection table alongside cadaveric dissection is preferable compared to being used alone.

### Limitations on the use of Anatomage in Anatomy Education

In this study, the inclusion of educational videos and animations to the Anatomage was emphasized by the students as a way of improving the equipment. Another point stated included the addition of more embryological specimens to the Anatomage.

## Data Availability

All data produced in the present study are available upon reasonable request to the authors

## Acknowledgment

The authors wants to acknowledge the Management of Edo State University Uzairue and the Medical Students that gave their consent for this work to be carried out.

